# Multi-component chlorination intervention to reduce neonatal infections in healthcare facilities in western Kenya (CLEAN Trial): study protocol for a cluster randomized controlled trial

**DOI:** 10.64898/2026.09.18.26363445

**Authors:** Yoshika S. Crider, Mwale Chiyenge, Erick Odoyo, Joyce Kisiangani, Jeremy Lowe, Josline Wangia, Blastus Bwire, Mwanzia Kioko, Kelly T. Alexander, Carol Nekesa, Lillian Musila, Phelgona Otieno, Amy J. Pickering

## Abstract

**Background:** The proportion of births occurring at healthcare facilities is rising globally, yet the birthing environment in healthcare facilities in low-income settings is often contaminated with bacterial pathogens, including antibiotic-resistant pathogens, that can lead to serious infections for newborns and their mothers. There is a need for effective strategies to reduce environmental contamination in healthcare facilities to reduce infection risks among facility-born neonates and their mothers.

**Methods:** We designed the CLEAN (<u>C</u>h<u>L</u>orine to reduce <u>E</u>nteric and <u>A</u>ntibiotic resistant infections in <u>N</u>eonates) cluster randomized controlled trial in western Kenya to evaluate the impact of a multi-component chlorination intervention on environmental contamination and maternal and neonatal infection risks. Thirty-six medium-sized public health facilities will be randomized in a 1:1 allocation ratio to receive a passive chlorination technology for water supply treatment paired with a reliable supply of chlorine-based disinfectant or status quo. Up to 22,500 mothers-neonate dyads will be enrolled and followed from birth through 28 days to collect symptoms of infection and mortality, with a subset of mother-neonate dyads selected for rectal swab collection to measure rectal colonization with sepsis-associated bacterial species. Environmental samples will be collected to measure bacterial pathogens on staff hands, high-touch surfaces, and in water supply. The primary objectives of the study are to evaluate the impact of the intervention on the following outcomes: (1) rectal carriage of bacterial pathogens one week post-birth among facility-born neonates and their mothers, (2) cumulative incidence in the first 7 days post-birth of possible serious bacterial infection among facility-born neonates, and (3) cumulative incidence in the first 7 days post-birth of symptoms of possible maternal sepsis.

**Discussion:** This study will generate evidence on the effectiveness of a novel chlorination intervention to reduce healthcare associated infections, including antibiotic resistant infections, and improve maternal and neonatal survival.

**Trial registration:** Clinical Trials NCT06824350. Registered 7 February 2025, https://clinicaltrials.gov/study/NCT06824350

## Background

The neonatal period is critical for child survival, as nearly half of deaths globally among children under 5 occur in the first month of life (1). Skilled birth attendance can substantially reduce maternal and neonatal mortality by ensuring access to medical care and resources (2), and the “proportion of births attended by skilled health personnel” is one of two indicators of progress toward reducing maternal mortality in the Sustainable Development Goal Agenda (3). Globally, the proportion of births occurring at health facilities has increased over time (4). However, exposure to health facilities has also been found to increase neonatal colonization with bacterial pathogens, including multidrug resistant pathogens, a precursor to infections (5,6). Serious bacterial infections are a leading cause of neonatal mortality globally, and especially in Sub Saharan Africa (SSA) (7), where an estimated 270,000 neonates die of sepsis annually (8).

Horizontal transmission (i.e., from the hospital environment) has been implicated in low-and middle-income countries (LMICs) as a dominant cause of neonatal colonization with antibiotic resistant bacteria (9). These include the ESKAPE pathogens (*Enterococcus faecium*, *Staphylococcus aureus*, *Klebsiella pneumoniae*, *Acinetobacter baumannii*, *Pseudomonas aeruginosa* and *Enterobacter* spp), which cause the majority of hospital-acquired infections and are increasingly resistant to multiple classes of antibiotics (10). Several of these bacteria have been identified as major causes of neonatal sepsis (11), and studies have documented the rapid gut and nasal colonization of both mothers and neonates with antibiotic resistant ESKAPE pathogens in HCF settings (5,6). A study in Bangladesh found that after a median hospital stay of three days following childbirth, maternal rectal colonization with carbapenem-resistant organisms increased from 13% to over 70% (5). A study in Kenya found that 55% of neonates admitted to a county referral hospital rapidly acquired extended-spectrum beta-lactamase-producing (ESBL) *Enterobacterales* (6). Neonatal gut colonization with beta-lactamase genes is strongly associated with a subsequent clinical sepsis diagnosis (12), suggesting that rectal colonization with antibiotic resistant bacteria is an important precursor to sepsis and that preventing this colonization may reduce sepsis incidence.

Preventing exposures to antibiotic resistant pathogens is especially crucial because of increasing resistance to first-line antibiotics for treatment. For example, in LMIC settings where advanced treatment options and laboratories for confirmatory testing are often unavailable and inaccessible, current World Health Organization (WHO) guidelines for the treatment of neonatal sepsis include empirical treatment with ampicillin and gentamicin (13). A multi-country study of antimicrobial resistance in neonatal sepsis identified resistance to ampicillin and gentamicin in 60% of Gram-negative bacteria isolates from confirmed sepsis cases.(12) This complicates treatment, results in longer hospital stays, and increases healthcare costs. Growing antibiotic resistance will add to an already expensive healthcare burden; in SSA, neonatal sepsis is estimated to cost up to almost $500 billion in economic losses annually (14).

A hygienic environment, including safe water and clean surfaces, is fundamental for infection prevention and control (IPC) (15). Contaminated water supplies and surfaces and healthcare worker hands have been implicated in the spread of healthcare associated infections across settings (15–17), and a recent study of HCFs in 14 LMICs found that only 31% had access to a water source free from *E. coli* contamination (18). A 2016 evaluation of water quality at HCFs in Kenya’s Siaya County by the non-governmental organization CARE-Kenya found that 50% (13/26) of source water samples were contaminated with *E. coli*. (19) and only 29.5% (13/44) of functional drinking water stations in HCFs had detectable free chlorine residual to protect water quality during storage (20). While hygiene conditions in health facilities are understood to be linked to infections, there is limited evidence on effective interventions. A 2019 systematic review of health facility-based water, sanitation, and hygiene (WaSH) interventions to reduce nosocomial infections identified only 3 eligible, high quality studies, but all reduced healthcare associated infections (21). Thus, there is a clear need to develop and generate evidence for the effectiveness of practical interventions to improve health facility conditions and prevent healthcare associated infections among vulnerable patient populations.

Our team has designed a multi-component chlorination intervention that pairs water treatment via in-line chlorination devices, which can be installed in existing infrastructure and operate without electricity, with a reliable supply of sodium hypochlorite, which is used for both surface disinfection and refilling the water treatment device for use in maternity units. This manuscript follows the Standard Protocol Items: Recommendations for Interventional Trials (SPIRIT) 2025 checklist (22) to describe the protocol for the CLEAN (<u>C</u>h<u>L</u>orine to reduce <u>E</u>nteric and <u>A</u>ntibiotic resistant infections in <u>N</u>eonates) cluster-randomized controlled trial that will evaluate the impact of this multi-component chlorination intervention in healthcare facilities on maternal and neonatal infections.

## METHODS AND ANALYSIS

### Study design

The CLEAN Trial is a parallel arm, cluster randomized controlled trial with a 1:1 allocation ratio. The health facility is the unit of randomization, and outcomes are measured in eligible individuals within health facilities. Intervention facilities will receive an in-line chlorination technology for water supply treatment and a reliable supply of sodium hypochlorite disinfectant. Both control and intervention facilities will receive infection prevention and control training. The primary objectives of the study are to evaluate the impact of the intervention on the following outcomes: (1) rectal carriage of bacterial pathogens one week post-birth among facility-born neonates and their mothers, (2) cumulative incidence in the first 7 days post-birth of possible serious bacterial infection among facility-born neonates, and (3) cumulative incidence in the first 7 days post-birth of symptoms of possible maternal sepsis. Secondary and additional outcomes include the intervention effects on bacterial contamination of water supply, on staff hands, and on high-touch surfaces in maternity wards, free and total chlorine concentrations in intervention facility water samples, and other outcomes described in **Table 1**. To assess the contribution of vertical versus horizontal transmission, both vaginal and rectal swabs will be collected from mothers who are enrolled prior to delivery. This study will provide practical insights into how to optimize and target facility-based WaSH interventions to reduce transmission of bacterial pathogens, including antibiotic resistant pathogens, to reduce healthcare-associated maternal and neonatal infections.

**Figure 1.**
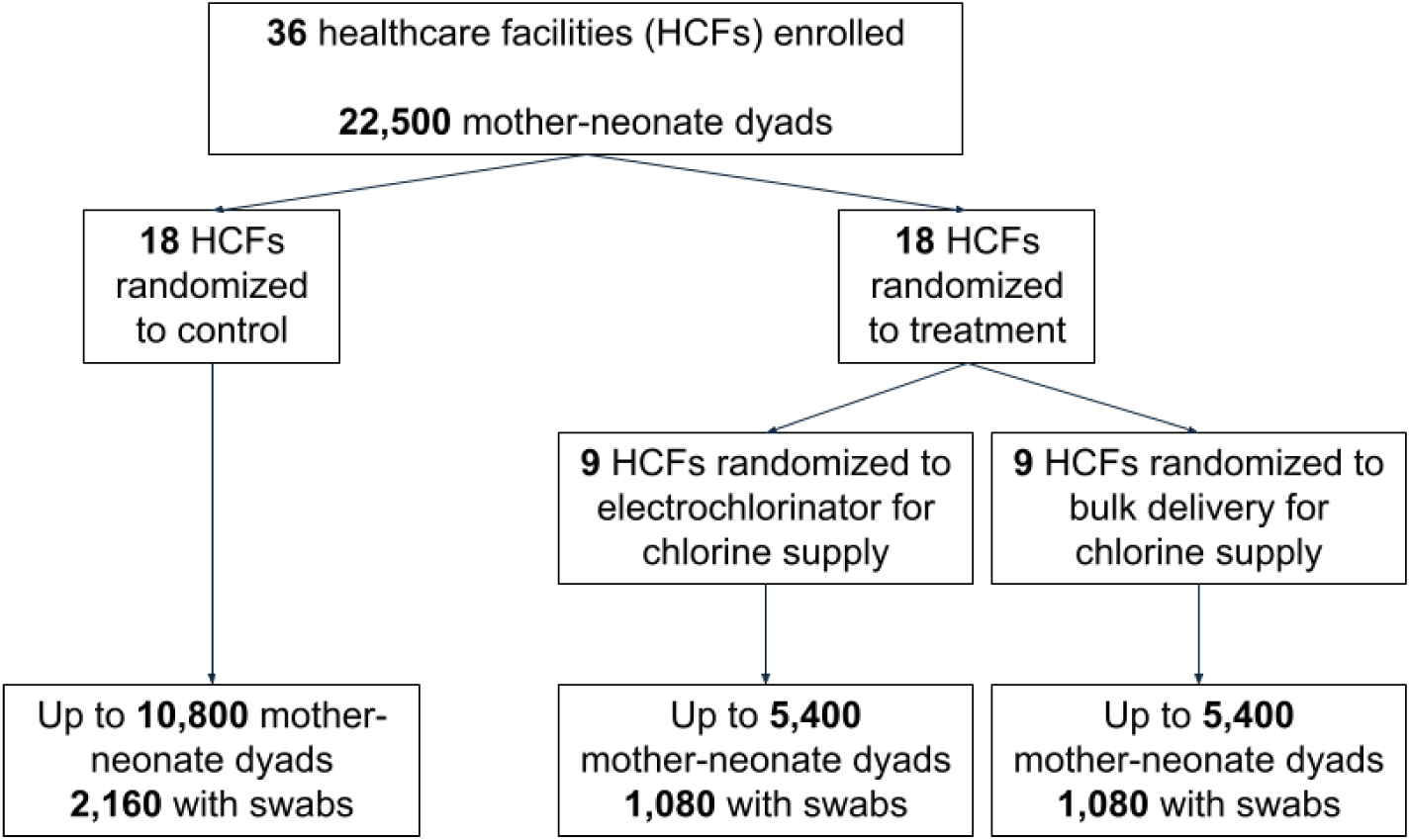
Participant flow chart.

**Figure 2.**
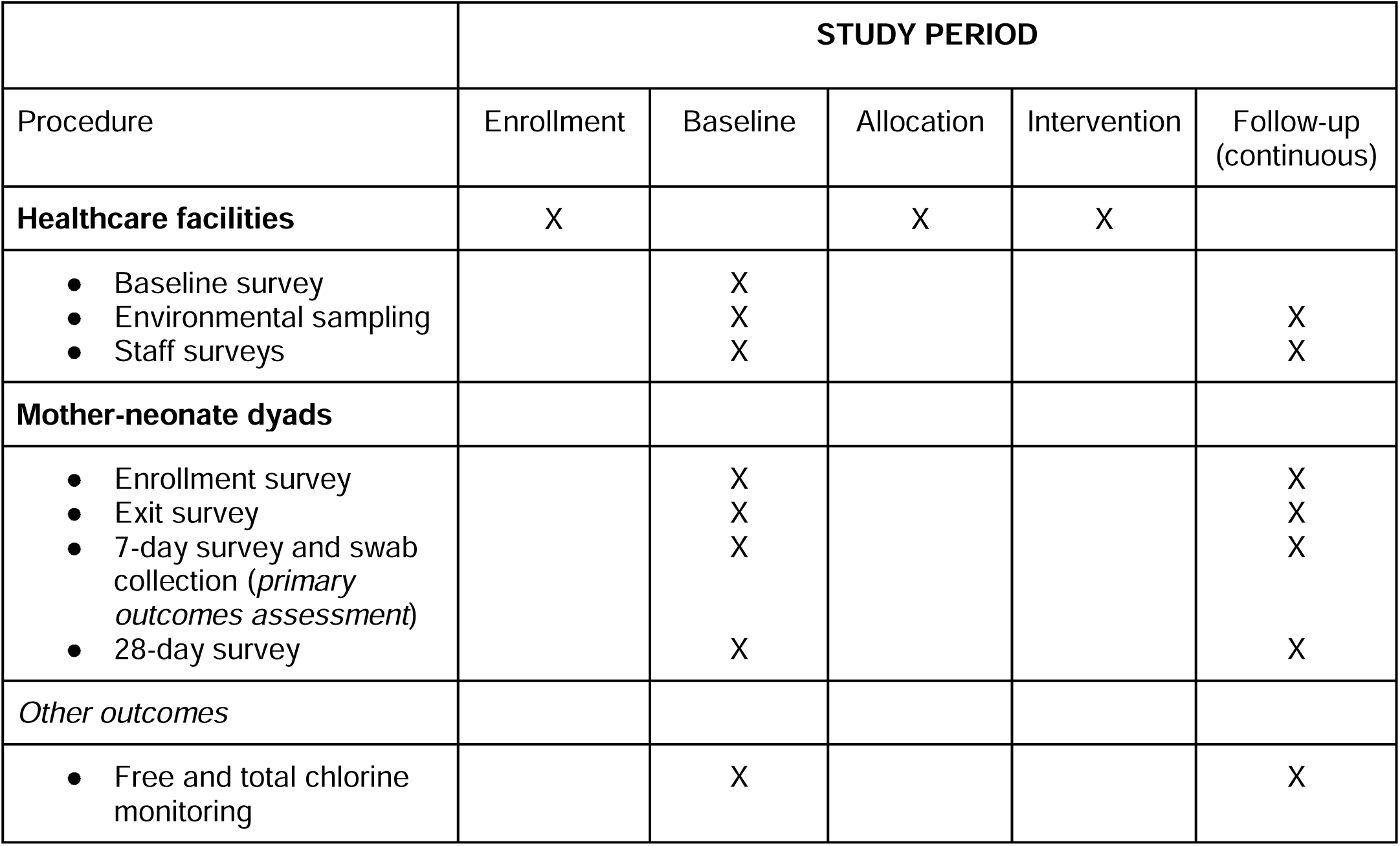
Schedule of enrollment, interventions, and data collection time points (SPIRIT Figure)

**Table 1.**
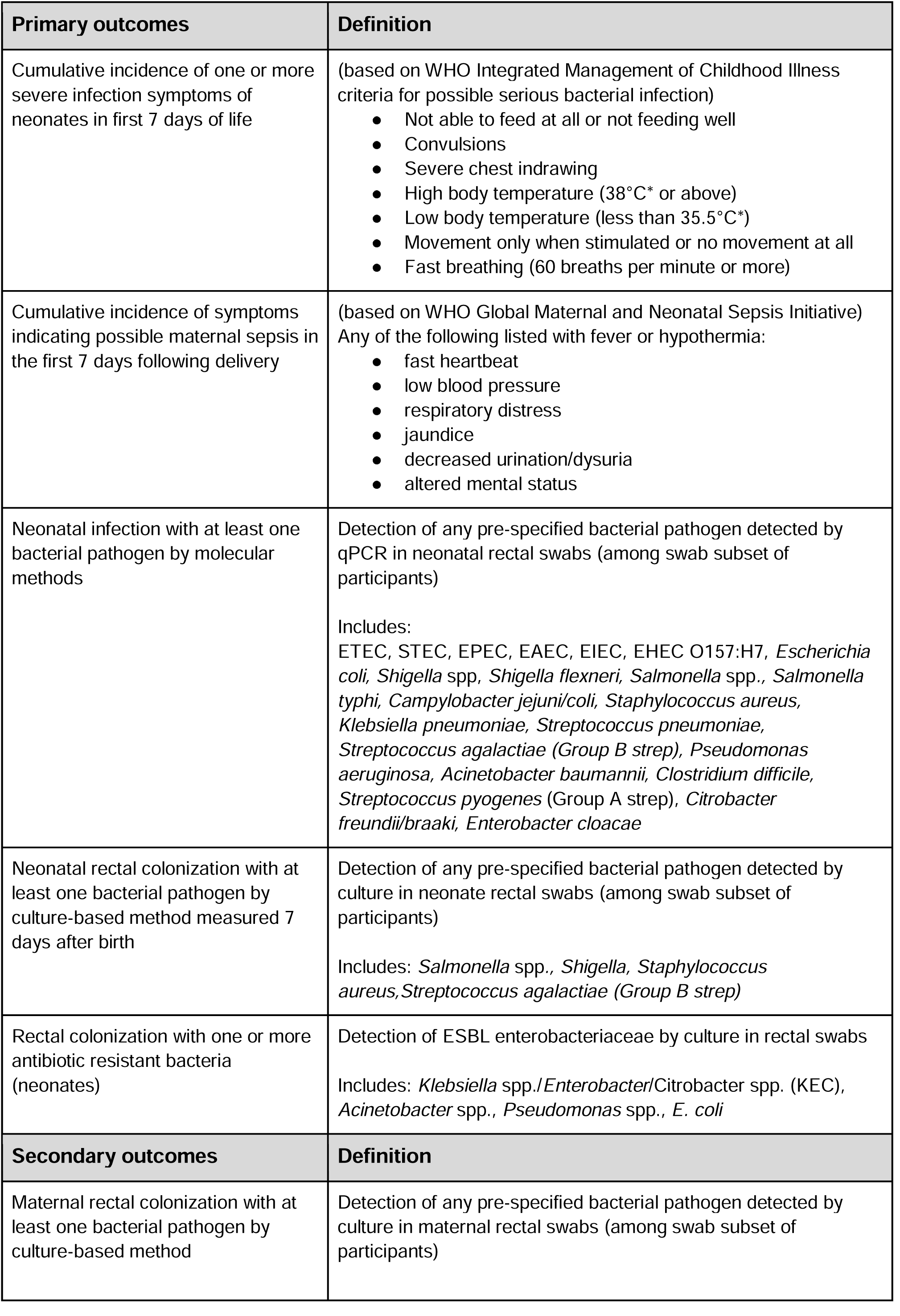
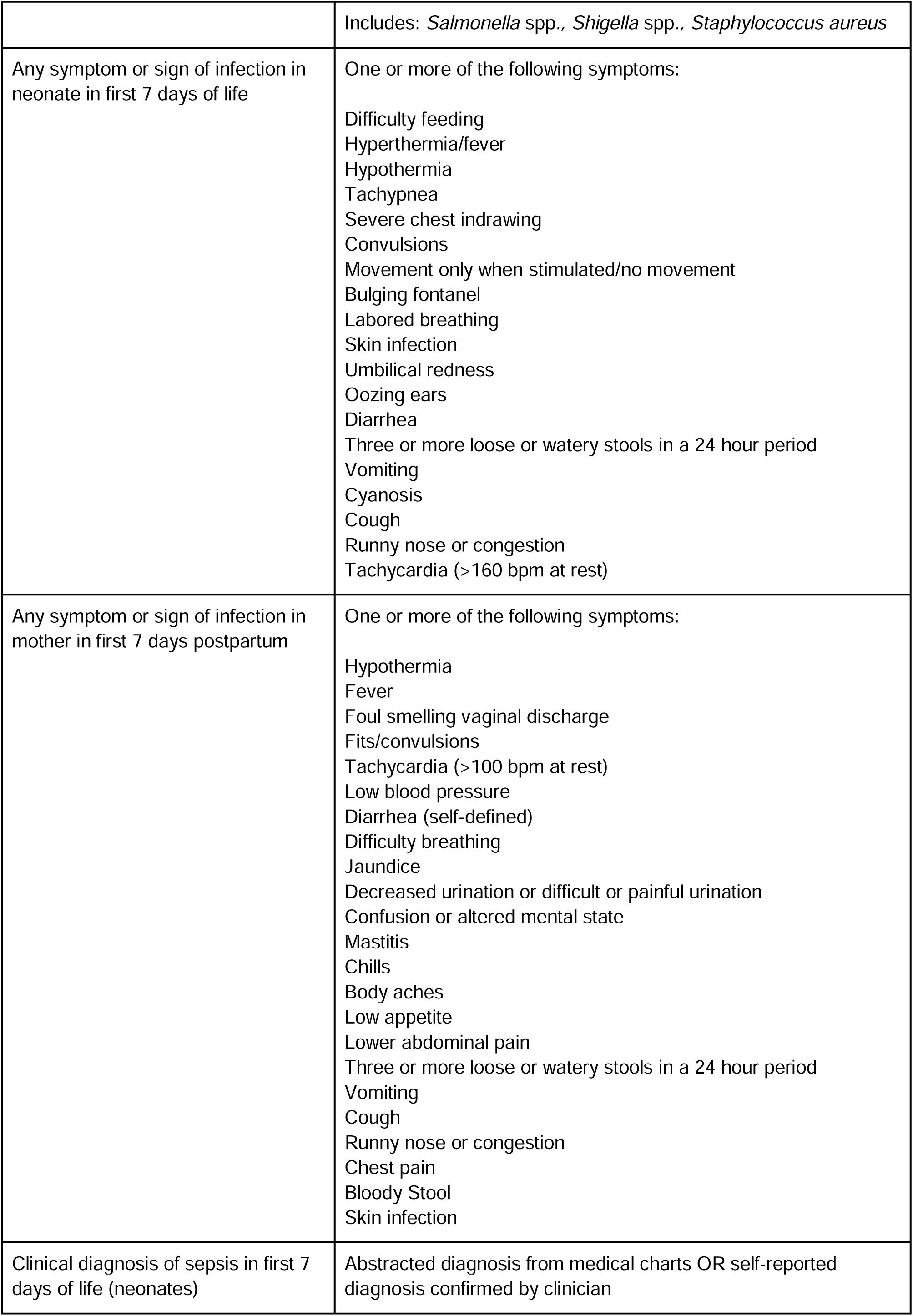
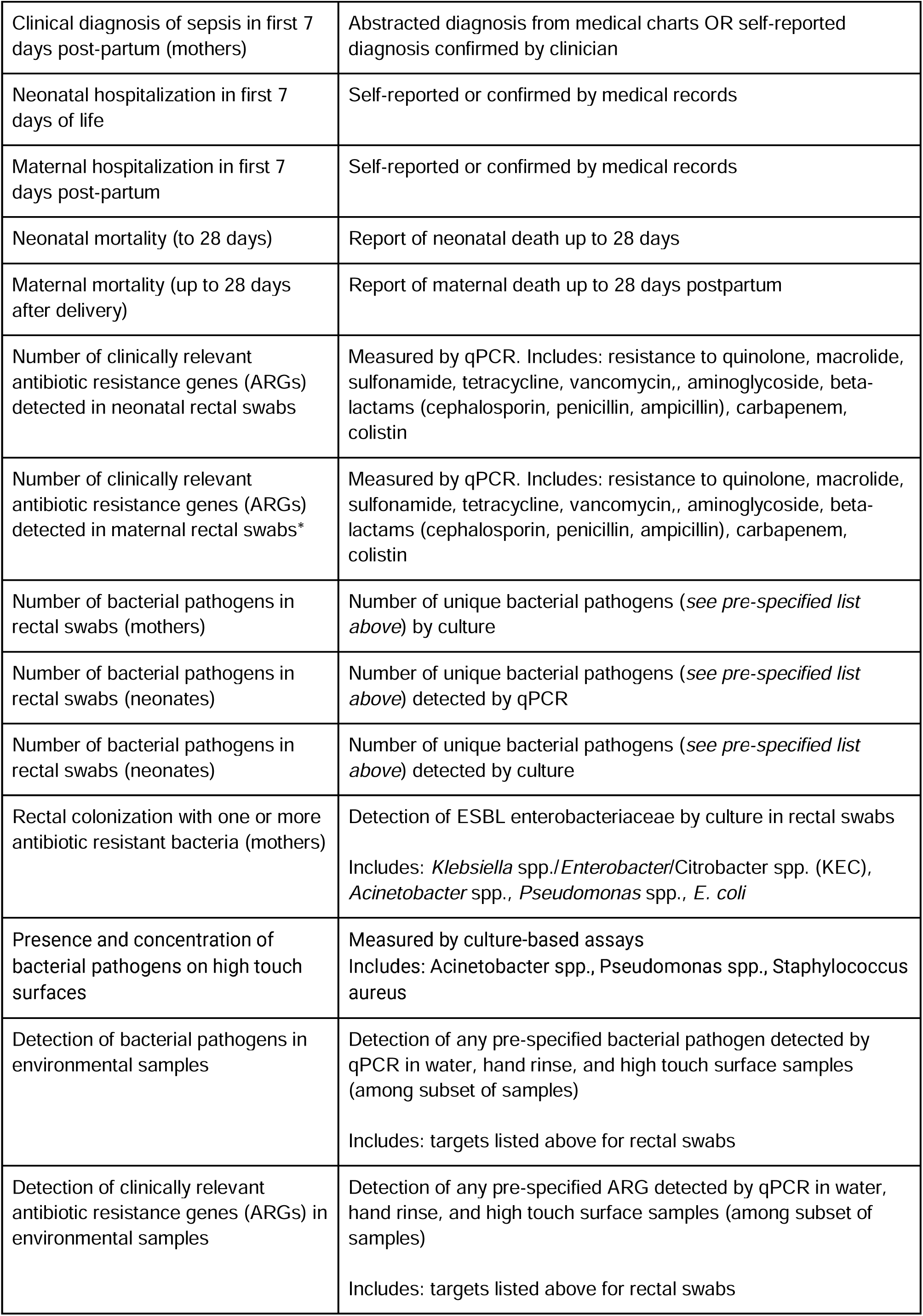
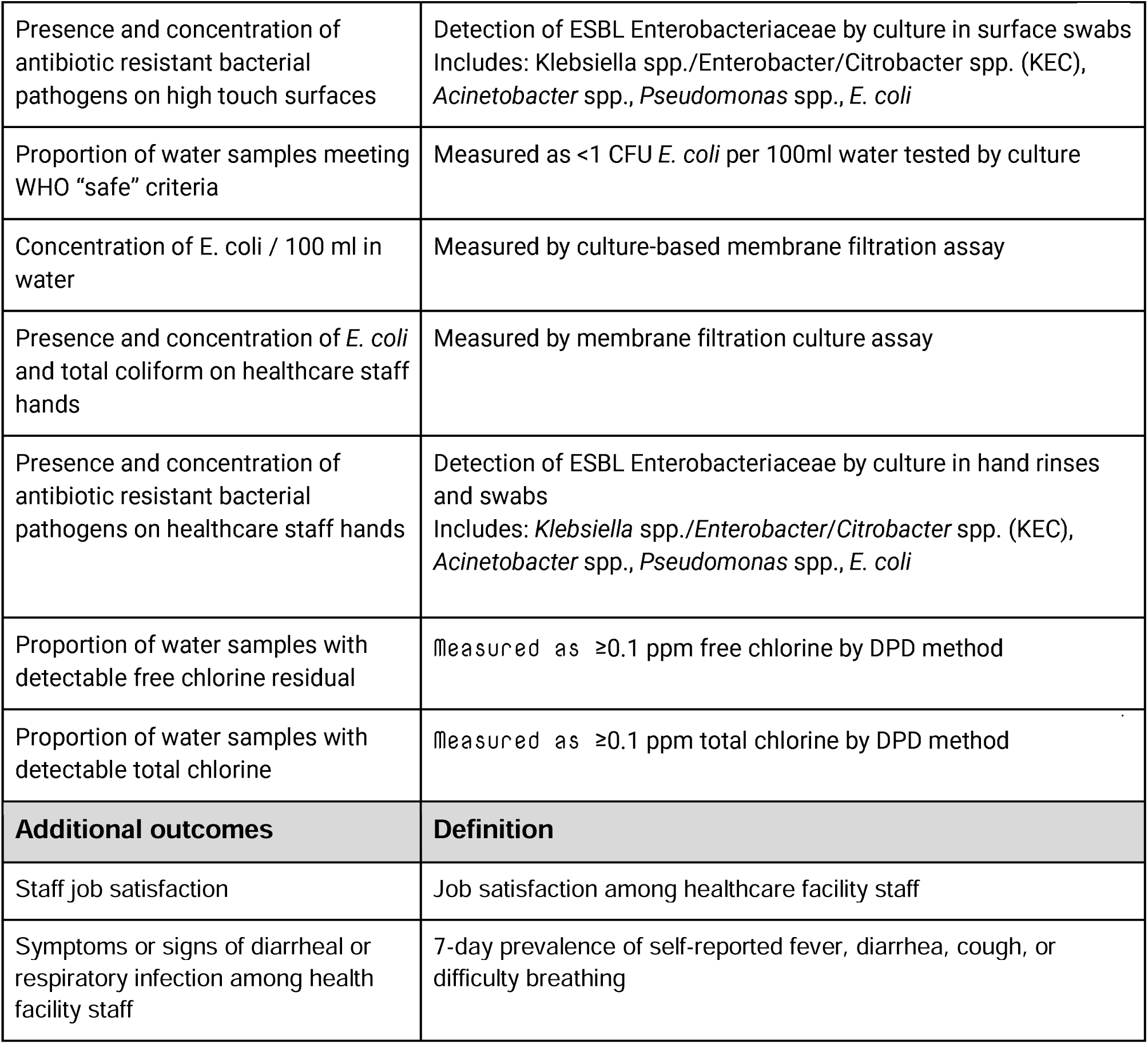
Study outcomes.

### Study site

The study is set in public healthcare facilities in Busia, Homa Bay, Kisumu, Migori, and Siaya counties in western Kenya.The main testing laboratory is located at the Kenya Medical Research Institute (KEMRI) in Kericho, Kenya, and additional molecular analysis will be performed at the University of California, Berkeley, California, USA.

**Figure 3.**
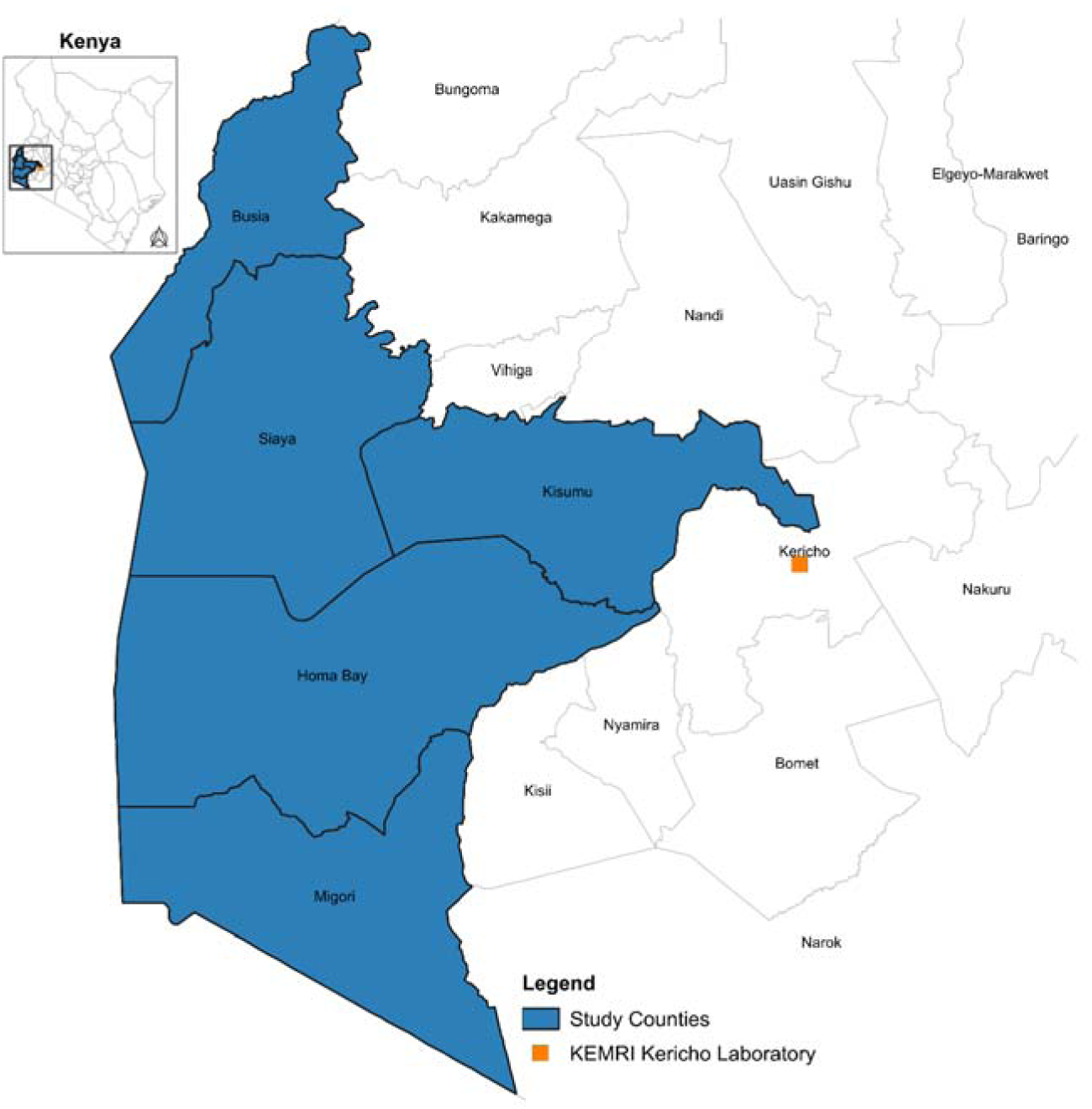
Study location.

### Eligibility criteria

#### (i) Criteria for inclusion and exclusion of study sites

Eligible health facilities are public facilities that meet the following criteria: (1) 25 or more live births per month on average; (2) no ongoing, consistent facility-level water treatment; (3) located within 3 hours of a study field office for sample transport logistics; and (4) compatible infrastructure and source water for in-line chlorination. In-line chlorination is compatible with infrastructure that includes the presence of water taps and centralized storage tanks on-site, with low turbidity (<5 NTU) and iron (<1.0 mg/L) concentrations in raw water supply.

#### (ii) Criteria for inclusion and exclusion of study participants

##### Mother/neonate dyads

All pregnant people (hereafter referred to as “mothers”) who arrive at enrolled facilities to deliver are screened for enrollment. Mothers under 15 years of age and those with significant medical challenges that prevent informed consent will be excluded from the study. Any decisions about medical fitness to consent will be made by healthcare providers. For participants selected for swab collection, additional criteria include living 2 hours or less from the health facility (to allow for a home visit) and no medical reason to disqualify vaginal (for mothers only) or rectal swab collection (for mothers and neonates). Any decisions about medical disqualification for swab collection will be made by healthcare providers. In the event of a multiple birth, all neonates may be enrolled. Mothers who deliver stillbirths may continue to participate because they will be similarly exposed to the health facility environment, however, stillbirths will be excluded from the neonatal analysis. Mothers who arrive with false labor and miscarriage (<28 weeks gestation) are excluded. Mothers who are enrolled at study facilities and subsequently referred to other facilities will continue to be followed for data collection.

##### Staff

All healthcare staff 18 years of age or older are eligible to participate in hand rinse sampling and workplace satisfaction surveys.

### Sample size

We determined the minimum detectable effect for two neonatal primary outcomes for infection (i.e., full sample “main cohort”) and gut carriage (i.e., swab subgroup) for a clustered design with binary outcomes.(23) We assume an intracluster correlation coefficient of 0.02 for outcomes within clusters (health facilities), based on prior research that found a median ICC of 0.019 (range = 0.0003-0.215) for outcome measures in maternal and perinatal health (24). We assume 80% power and a two-sided alpha=0.05. For our primary outcome of neonatal infection, we assume 22% of neonates in control facilities will report signs of serious infection, based on estimates of neonatal infection from hospital-based studies in LMIC settings ranging from 18% (25) to 22% (26). We calculate our minimum detectable effect assuming 18 health facilities per arm and 600 births per facility (assuming 25 births per facility per month), with 10% loss to follow up, allowing us to detect a 25% reduction in the outcome. For our primary outcome of neonatal gut carriage, we assume that 39% of neonates in control facilities will have at least one pre-specified bacterial enteropathogen, based on a prior study conducted by members of our team in northern Kenya that found 39% of neonatal stool samples were positive for the enteropathogen enteroaggregative *E. coli* (27). With 96 swab enrollments per facility (average 4 per facility per month), we are able to detect a 20% reduction in this outcome. We will target 5 enrollments per month on average, which allows for 20% respondent attrition in the swab subgroup. Our calculations assume one neonate enrolled per mother. However, in the event of multiple births, we will enroll all neonates. Mothers with stillbirths may continue to be followed in the study. In the event of a maternal death, neonate follow up will be discontinued.

### Enrollment

Informed consent will be obtained from all participants by study staff. Mothers will provide parental permission for rectal swab collection from their neonates. In the subset of mother/neonate dyads selected for swab collection, informed consent will include permission to retain deidentified swab specimen isolates indefinitely for further analysis. Participants are provided with a small token gift for each survey completed.

### Intervention

Prior to facility randomization, the lead infection prevention and control (IPC) staff member at every enrolled healthcare facility (control and intervention) will be invited to attend a study-sponsored centralized 1-day IPC refresher training. This refresher training will be led by an experienced trainer and focus on guidelines for safe water and surface disinfection for infection prevention and control in healthcare settings. Participants will be encouraged to provide peer mentoring to other IPC staff at their health facility. After facilities are randomized, health facilities assigned to the control group will receive no facility-level intervention, reflecting the status quo of water infrastructure and IPC practices in health facilities in Western Kenya.

Health facilities assigned to the intervention group will receive a multi-component facility-level intervention that includes:

1. One or more in-line water chlorination technologies for treatment of all water used in the maternity units, and;
2. Continuous supply of chlorine either a) through timely bulk delivery of liquid/powder chlorine for in-line chlorination refills and surface disinfection, or 2) an electrochlorinator technology for on-site liquid chlorine production. Health facilities will also receive necessary hardware to use the chlorine for cleaning purposes, including mops, buckets, spray bottles, and, for the bulk delivery facilities, 20L jerry can, measuring cup, and stirring rod for preparing dilute chlorine solution. A sensitization training will be provided at each intervention facility to provide instructions for hardware use and to identify IPC champions to support supervision and sustainability through cleaning staff turnover.

Facilities in the intervention group will be evenly randomized to the bulk delivery or electrochlorinator options in order to identify tradeoffs between the approaches for sustained disinfectant supply, to inform cost effective scaling of the intervention in the future. The intervention may be discontinued at the request of the health facility administrator-in-charge. There will be no changes or restrictions to patient care during the trial.

### Outcomes

The primary outcomes include (1) 7-day cumulative incidence of mother-reported symptoms of serious bacterial infection among neonates and possible sepsis among mothers in the week following birth and (2) prevalence of bacterial pathogens associated with sepsis in rectal swabs collected from neonates and mothers at 7-days, using both culture and molecular methods. A number of secondary and additional outcomes are pre-specified in **Table 1**.

### Implementation monitoring

Presence of chlorine Study staff will conduct monitoring surveys to measure free and total chlorine residual in tap and stored water samples at all facilities at least twice per month. Chlorine will be measured with a Hach Digital Colorimeter (Model DR300, Hach Company, Colorado, USA), which uses a DPD method (N,N-diethyl-p-phenylenediamine) to quantify free and total chlorine concentrations.

### Maintenance and repairs

Study staff will provide free repair and replacement for any broken intervention materials for the duration of the study.

### User uptake

All intervention facilities will receive a daily cleaning log with activities related to chlorine use in the maternity unit, in order to monitor use of chlorine for activities along our hypothesized causal pathway (e.g., preparation of chlorine dilutions, disinfection of high-touch surfaces). During intervention delivery, maternity unit cleaning staff will be instructed to fill the log daily.

### Cost tracking

Costs for all materials and labor for installation and repairs will be recorded. The volume of chlorine delivered (for bulk delivery intervention group) and produced (for electrochlorinator intervention group) will be monitored to calculate total costs associated with consumables. Study staff will record any materials and labor required for repairs to calculate total maintenance costs.

### Piloting

Study procedures were refined during a pilot phase of data collection in late 2024.

### Randomization

The facilities will be randomized in a 1:1 allocation ratio to receive the chlorination intervention or status quo. We will stratify by county and by facility size (based on monthly births), in order to ensure an even distribution of facility size (i.e., a cluster-level characteristic) between intervention and control groups. We will order facilities by descending size and randomize within each block of 2 facilities from the largest to smallest facility. For any county with an odd number of facilities, the smallest county will be included in the randomization of the next closest county with an odd number of facilities. A member of the research team will randomly select one healthcare facility to receive the intervention within each block using a random number generator in R. Half of intervention facilities will be assigned to receive an electrochlorinator (9 total). Each county will be assigned a supply of electrochlorinators equal to half the number of intervention facilities, rounded down. We will pair nearest counties that have an odd number of intervention facilities and randomly select which of the two counties receives the additional electrochlorinator. Within counties, intervention facilities will be randomly selected for the available electrochlorinators. The remaining intervention facilities will receive bulk chlorine deliveries.

### Facility surveys

All surveys will be conducted on electronic tablets using SurveyCTO software.

#### Baseline health facility assessment

Prior to implementing the intervention, we will conduct a baseline survey with environmental sampling at all health facilities. The baseline survey will be conducted with multiple health facility staff members with expertise in the facility’s infection prevention and control practices, water and electricity infrastructure, maternity unit resources and services, and environmental cleaning practices.

**Figure 4.**
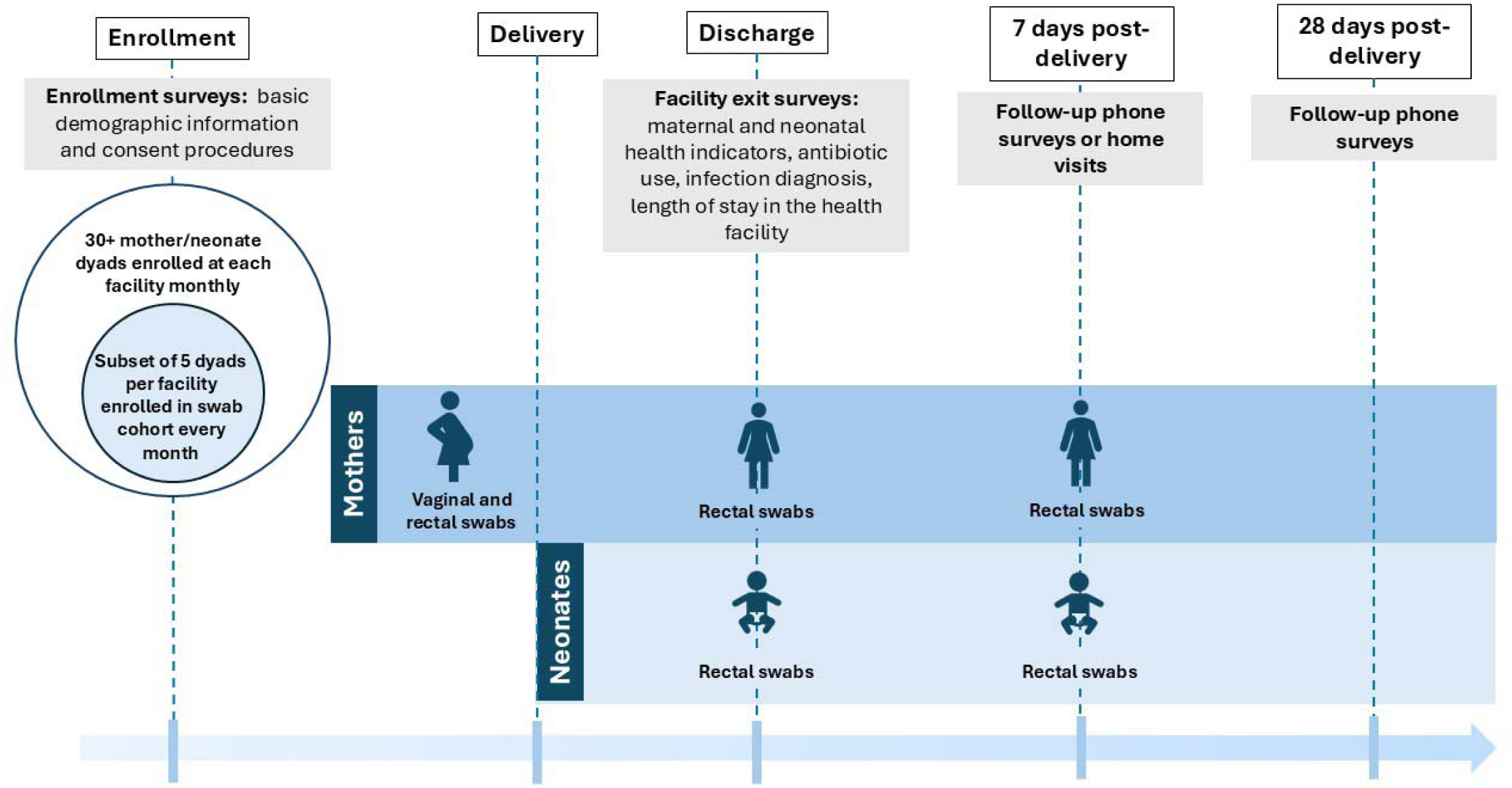
Timeline of study activities.

### Mother surveys

#### Time point 1: Enrollment survey

Mothers who are enrolled at health facilities prior to delivery will complete a brief enrollment survey to collect basic demographic information and consent procedures. For those selected for the swab group, vaginal and rectal swabs will be collected by trained study staff.

#### Time point 2: Mother facility exit survey

Following delivery and prior to discharge from health facilities, all enrolled mothers will complete a facility exit survey conducted by trained staff. Mothers are also eligible to be enrolled at this time point, using an enrollment module within the facility exit survey. During the facility exit survey, trained staff will collect data on birth outcomes, birthweight, antibiotic use since hospital admission, and any infection diagnosis made at the health facility. Additional data abstraction from medical records to record treatment and clinical diagnoses will be conducted by trained study staff. Mothers will be provided a handout describing neonatal signs of serious infection and requested to return to the facility for medical care if they observe any of the following signs and symptoms in their neonate: difficulty feeding, too hot or too cold, convulsions, yellow palms or soles of feet, little movement, chest indrawing, or fast breathing. Contact information will be collected from the mother, including up to 2 alternate phone numbers for household or community contacts, to ensure the mother can be reached in 7 days. For those selected for the swab group, rectal swabs are collected by trained study staff from both the mother and the neonate. Additional contact information will be collected from mothers in the swab group to facilitate a home visit (e.g., recording landmarks near their home).

#### Time point 3: Mother 7-day survey

At 7 days following birth, trained study staff will contact mothers to record specific self-reported signs and symptoms of infection in themselves and their neonates. The majority of mothers will be contacted by phone; the subset of mothers who provide swab samples will receive a home visit. For neonates, symptoms of possible serious bacterial infection, as defined by guidelines for Integrated Management of Childhood Illness include: inability or refusal to breastfeed, convulsions, fast breathing (60+ breaths per minute), chest in drawing during breathing, elevated temp (fever), low temperature (hypothermia), movement only when stimulated/no movement.(13) We will additionally record the following: irritability, high pitched crying, bulging fontanel, labored breathing, jaundice, skin pustules, umbilical redness, oozing ears, diarrhea, vomiting, cyanosis or death. Neonates with reported symptoms indicative of serious infection will be referred back to the healthcare facility for treatment. If a death is observed, date and official cause of death will be recorded. If no official cause of death is available, the World Health Organization 2022 verbal autopsy checklist will be used to ascertain likely cause of death.(28) There is no standard definition for maternal possible serious bacterial infection (29,30), so we use symptoms identified by the World Health Organization’s Global Maternal and Neonatal Sepsis Initiative as suggestive of maternal sepsis.(31) Symptoms include any of the following listed with fever or hypothermia: fast heartbeat/low blood pressure, respiratory distress, jaundice, decreased urination/dysuria, or altered mental status. Mothers with these symptoms will be requested to report back to the health facility immediately. Additionally, we will record the following: general discomfort, chills, body aches, loss of appetite, lower abdominal pain, foul smelling vaginal discharge, headaches, diarrhea, vomiting, mastitis, cough, and chest pain. Specifically, mothers will be asked: “Since you gave birth, has your newborn had [symptom name]?” for any individual symptom listed above. As a secondary outcome, mothers will be asked: “Since you gave birth, have you had [symptom name]?” for any individual symptom listed above. Survey staff will attempt to call the mother up to 3 times to collect this information, and alternate contact numbers collected at the exit survey will be used if a mother is unreachable.

#### Time point 4: 28-day survey

At 28 days following birth, trained staff will attempt to contact all mothers by phone to record any maternal or neonatal deaths. Alternate contact numbers collected at the exit and/or 7-day surveys may be used to reach mothers.

### Verbal autopsy

Trained staff will conduct a verbal autopsy survey following any maternal or neonatal death recorded during the study period. Verbal autopsies for neonatal deaths will be conducted with the mother. Verbal autopsies for maternal deaths will be conducted with the closest family member. These may be conducted in person or by phone and conditional on the verbal consent of the respondent.

### Sample collection

#### Vaginal swab collection

We will collect rectal and vaginal swabs from mothers who are selected for swab sampling and enrolled prior to delivery (at time point 1) to assess vaginal and rectal carriage of bacterial pathogens prior to giving birth. These swabs will be analyzed using molecular-and culture-based methods, in order to assess potential vertical transmission of pathogens from mother to neonate during birth.

#### Rectal swab collection

We will collect rectal swabs from neonates and mothers after delivery (before hospital discharge), and 7 days after birth to assess rectal carriage of bacterial pathogens previously linked to neonatal infections, hospital-acquired infections, and sepsis (11,12,32), including antibiotic resistant bacteria, using molecular-and culture-based methods.

### Environmental sampling: hands, water, and surfaces

We will conduct longitudinal environmental sampling to assess bacterial contamination, including with antibiotic resistance, in maternity unit environments (water supply and high-touch surfaces) and healthcare provider hands. We will sample prior to intervention (baseline) and after intervention delivery (follow-up) every 4 months over the 2-year study period. During the baseline visit, trained staff will identify up to eight high touch surfaces within maternity units; these same surfaces will be consistently sampled at each time point. Two healthcare providers and one custodial staff member working in the maternity unit at the time of the sampling visit will be selected for hand rinse sampling. Up to four water samples will be collected from the maternity ward. A subset of environmental samples will be additionally processed via the molecular methods for rectal swabs described below. All environmental samples will be processed at the KEMRI Kericho laboratory, with additional molecular analysis conducted at UC Berkeley.

### Laboratory assays

#### Culture assays

*Environmental samples.* Water samples and hand rinse samples will be processed via membrane filtration and evaluated for *E. coli*, total coliform concentrations (CHROMagar ECC; EF323), and ESBL-producing *Enterobacteriaceae*, *Acinetobacter* and *Pseudomonas* presence (CHROMagar ESBL; ESRT2). 100 µl of environmental swab sample suspensions (Puritan 25-83004 PD NB) will be plated on sheep blood agar to quantify bacterial loads and on chromogenic media to qualitatively detect the presence of target organisms.

#### Vaginal and rectal swab samples

Swab sample suspensions (Copan FecalSwab with Cary Blair medium; 4C028S) will be streaked onto chromogenic differential or selective media using 10 µl inoculating loops to qualitatively assess the presence of target enteric and sepsis-associated bacteria.

#### Detection of target organisms

Selective or differential media will be used for the qualitative isolation and detection of target organisms as indicated in **Table 2**. For the majority of the ESKAPE-E target isolates, the inoculated plates will be incubated aerobically at 37 °C for 24 - 48 h and identified by observing the typical growth characteristics on the respective chromogenic agar and the selective/differential media. A subset of isolates will be analyzed with MALDI-TOF mass spectrometry (<u>MALDI Biotyper® system</u>, Bruker).

**Table 2.** Culture media for isolation and detection of target organisms in water, hand rinse, surface swab, and rectal and vaginal swab samples.

| Organism | Selective/differential detection media | Samples processed |
| --- | --- | --- |
| *Staphylococcus aureus* | CHROMagar *Staph aureus* (TA672) | Surface swabs, rectal and vaginal swabs |
| Group B *Streptococcus* | CHROMagar StrepB (SB282) | Rectal and vaginal swabs |
| *Salmonella* species | Oxoid SS Agar Modified (Thermo Scientific CM0533B) |  |
| *Shigella* species | Oxoid SS Agar Modified (Thermo Scientific CM0533B) |  |
| ESBL *Enterobacteriaceae*<br>(*E. coli*, *Klebsiella* spp.,<br>*Enterobacter* spp.,<br>*Citrobacter* spp.) | CHROMagar ESBL (ESRT2) | Water, hand rinses, surface<br>swabs, rectal and vaginal<br>swabs |
| ESBL *Pseudomonas* | CHROMagar ESBL (ESRT2) |  |
| ESBL *Acinetobacter* | CHROMagar ESBL (ESRT2) |  |

#### Molecular assays

*Rectal swab samples.* Swab samples collected from neonates (preserved in Zymo DNA/RNA shield) will undergo DNA extraction using the Quick-DNA Fecal/Soil Microbe Microprep kit protocol (Zymo D6012) or similar, and the extracted DNA will be analyzed for the presence of bacterial pathogens and antibiotic resistance genes (ARG) of clinical relevance using custom qPCR TaqMan Array Cards (TAC) whose microfluidic design enables 48 qPCR assays to be performed simultaneously (Applied Biosystems). The custom TAC will include assays for enteric and HAI-associated bacterial pathogen gene targets (including diarrheagenic *Escherichia coli, Shigella* spp., *Salmonella* spp*., Campylobacter jejuni/coli, Clostridium difficile, Staphylococcus aureus, Klebsiella pneumoniae, Streptococcus pneumoniae, Streptococcus agalactiae (Group B strep), Pseudomonas aeruginosa, Acinetobacter baumannii, Streptococcus pyogenes* (Group A strep), *Citrobacter freundii/braaki, Enterobacter cloacae*), and ARGs conferring resistance to quinolones, macrolides, aminoglycosides, teracyclines, vancomycin, penicillins, cephalosporins, and last-resort antibiotics (colistin and carbapenems). The card will also include an endogenous control assay for bacterial 16S and an assay for a *Bacillus atrophaeus* internal extraction control spike-in.

### Assignment of interventions: Blinding

Health facilities and participants will not be blinded to the intervention, as this would be infeasible for an infrastructure intervention. Laboratory technicians at KEMRI who process samples will be blinded to intervention assignment. Principal Investigators and members of the research team who are involved in the chlorinator technology installations will not be blinded to intervention assignment. Principal investigators will be blinded to primary outcomes by treatment status until the end of the trial.

## Data collection and management

### Plans for assessment and collection of outcomes

Study staff will undergo intensive training on the survey instruments. No interim analyses are planned for primary outcomes. Mortality and other serious adverse events will be continuously monitored and reported as required by institutional IRBs and the study’s data and safety monitoring committee (DSMC). Surveys will be made publicly available along with deidentified datasets and replication code upon publication of results.

### Data management

All hospital, patient, and laboratory data will be collected on electronic tablets using SurveyCTO software.

### Confidentiality

Each participant is assigned a unique study identification code to link encrypted survey data and samples. De-identified datasets will be made publicly available following publication of results.

### Statistical methods

Analyses will be intent-to-treat. Any mother who delivers in a facility assigned to the intervention will be analyzed in the intervention group. We will combine the two intervention arms (on-site chlorine production with electrochlorinator and chlorine delivery) into a single group. Our primary outcomes are binary indicators of infection and colonization. To compare intervention and control groups, we will estimate interpretable risk ratios using generalized linear models using a log-binomial method (family=binomial, link=log). If a log-binomial model does not converge, we will use a modified Poisson method (family=Poisson, link=log) to estimate risk ratios (33). If our baseline assumptions are incorrect and the outcome is rare (<10%), we will consider alternative model approaches. Models will include county, randomization block, and month fixed effects. In secondary covariate-adjusted analysis, we will report results of adjusted models that adjust for each of the following categorical covariates that has a prevalence of at least 5% and is associated with the outcome (likelihood ratio test p-value < 0.1): number of people in the household, reported food insecurity, asset index, child birth order (first-born or not), mother age, and mother educational attainment.

We will conduct subgroup analyses for neonate outcomes stratified by birthweight (normal and low (<2500 grams) and gestational age (term and preterm (<37 weeks)), reported exclusive breastfeeding, and neonate sex, as these are known to be associated with risk of neonatal illness. We will conduct a subgroup analysis for mother outcomes for primiparous versus multiparous status mothers. We will conduct subgroup analyses for all primary outcomes by season (wet vs dry), presence of improved water versus unimproved at the household, and presence of improved sanitation versus unimproved at the household.

### Interim analyses

No stopping rules are pre-specified for the study. A summary of serious adverse events, including mortality, by study arm will be prepared by a non-PI member of the research team and shared with the members of the data and safety monitoring committee (DSMC) at an annual meeting. No other members of the research team will view outcomes by study arm prior to the final analysis. This committee will have authority to stop the study if there is any evidence of harm to participants, which will be determined by their expert opinion.

### Missing data and loss to follow up

We will report loss to follow up by study arm. We define loss to follow up as any enrolled respondent for whom there is incomplete information on relevant primary outcomes. This includes respondents who are unreachable, who decline to participate after initial enrollment, or whose death is recorded before ascertaining 7 days endpoints.

If individual participant loss to follow up (at 7 days) is >20%, we will consider using inverse probability weighting using enrollment data. Additionally, if loss to follow up is >20%, we will assess the possible resulting bias to our estimates using 2 approaches. First, we will estimate the worst case scenarios using Manski bounds, where missing binary values are either all 1 or all 0. Second, we will estimate a more likely scenario by imputing sample means for missing values. Additionally, we will compare enrollment characteristics for respondents with complete cases versus respondents who were lost to follow up, in order to assess whether lost respondents may be systematically different from those who provided complete follow up data (i.e., if a missing at random assumption may not be appropriate).

### Oversight and monitoring

The trial has a 5-member data safety and monitoring committee (DSMC) that includes experts in clinical trials, maternal and neonatal health, biostatistics, and epidemiology. The committee will meet on an annual basis to review trial progress. If any member can no longer serve on the committee, a replacement will be appointed by the study PIs. The charter is available upon request. Members of the committee have research funding from the National Institutes of Health, the primary funder of this trial.

### Trial status

A pilot study was conducted at non-study facilities in 2024. Health facility enrollment began in late 2024, with participant recruitment beginning in February 2025.

## Declarations

### Ethics approval and consent to participate

We obtained ethical approval from the University of California, Berkeley, and the Kenya Medical Research Institute (SERU #5188), the primary sponsors of this trial. The University of Minnesota provided institutional approval via a formal IRB reliance agreement with UC Berkeley. Additionally, we obtained memorandums of understanding with county governments in all study site counties and are licensed by the National Commission for Science Technology and Innovation (NACOSTI/P/25/40230). Voluntary, informed consent is obtained from all adult and mature minor participants and parental permission is obtained for all neonate participants.

## Data Availability

This is a study design paper.

## Acknowledgements

We would like to acknowledge the County Health officials, hospital administrators and staff at the healthcare facilities enrolled in the study.

## Authors’ contributions

The trial protocol was drafted by YC, MC, EO, LM, PO, and AP. JK, JL, JW, BB, MK, RM, SO, KA, and CN contributed input. All authors read and approved the final manuscript.

## Availability of data and materials

De-identified dataset, surveys, and replication code will be made publicly available upon publication of results. The statistical analysis plan for this trial is available upon request to the corresponding authors. The DSMC charter is available upon request to the corresponding authors. Results will be disseminated in open access publications and via stakeholder meetings.

## Consent for publication

All authors approved the published manuscript.

