## Supplemental Information for "Multi-component chlorination intervention to reduce neonatal infections in healthcare facilities in western Kenya (CLEAN Trial): study protocol for a cluster randomized controlled trial"

### **SPIRIT Checklist for *Trials***

Complete this checklist by entering the page and line numbers where each of the items listed below can be found in your manuscript.

Your manuscript may not currently address all the items on the checklist. Please modify your text to include the missing information. If you are certain that an item does not apply, please state "n/a" and provide a short explanation. **Leaving an item blank or stating “n/a” without an explanation will lead to your manuscript being returned before review.**

Upload your completed checklist as an additional file when you submit to *Trials*. You must reference this additional file in the main text of your protocol submission. The completed SPIRIT figure must be included within the main body of the protocol text and can be downloaded here: <http://www.spirit-statement.org/schedule-of-enrolment-interventions-and-assessments/>

In your methods section, please state that you used the SPIRIT reporting guidelines, and cite them as:

Chan A-W, Tetzlaff JM, Gøtzsche PC, Altman DG, Mann H, Berlin J, Dickersin K, Hróbjartsson A, Schulz KF, Parulekar WR, Krleža-Jerić K, Laupacis A, Moher D. SPIRIT 2013 Explanation and Elaboration: Guidance for protocols of clinical trials. BMJ. 2013;346:e7586

|  |  | **Reporting Item** | **Page and Line Number** | **Reason if not applicable** |
| --- | --- | --- | --- | --- |
| **Administrative information** | | | | |
| Title | [#1](https://www.goodreports.org/reporting-checklists/spirit/info/#1) | Descriptive title identifying the study design, population, interventions, and, if applicable, trial acronym | Page 1  Line 1-2 |  |
| Trial registration | [#2a](https://www.goodreports.org/reporting-checklists/spirit/info/#2a) | Trial identifier and registry name. If not yet registered, name of intended registry | Page 2  Line 53-54 |  |
| Trial registration: data set | [#2b](https://www.goodreports.org/reporting-checklists/spirit/info/#2b) | All items from the World Health Organization Trial Registration Data Set | TRDS 1 (**Primary Registry and Trial Identifying Number): Page 2, Line 53-54**  TRDS 2 (**Date of Registration in Primary Registry): Page 2, Line 53**  TRDS 3 (**Secondary Identifying Numbers): NA**  TRDS 4 (**Source(s) of Monetary or Material Support): Pages 24-25, Lines 502-505**  TRDS 5 (**Primary Sponsor): Page 24, Lines 495-496**  TRDS 6 (**Secondary Sponsor(s)): NA**  TRDS 7 (**Contact for Public Queries): Page 1, Lines 27-28**  TRDS 8 (**Contact for Scientific Queries): Page 1, Lines 27-28**  TRDS 9 (**Public Title): Page 1, Lines 1-2**  TRDS 10 (**Scientific Title): Page 1, Lines 1-2**  TRDS 11 (**Countries of Recruitment): Pages 6-7, Lines 147-151**  TRDS 12 (**Health Condition(s) or Problem(s) Studied): Page 10, Lines 235-240**  TRDS 13 (**Intervention(s)): Pages 9-10, Lines 209-234**  TRDS 14 (**Key Inclusion and Exclusion Criteria): Pages 7-8, Lines 156-181**  TRDS 15 (**Study Type): Page 5, Lines 123-139**  TRDS 16 (**Date of First Enrollment)**  TRDS 17 (**Sample Size): Pages 8-9, Lines 183-202**  TRDS 18 (**Recruitment Status): Page 24, Lines 489-491**  TRDS 19 (**Primary Outcome(s)): Page 10, Lines 235-240**  TRDS 20 (**Key Secondary Outcomes): Pages 11-14 (Table 1)**  TRDS 21 (**Ethics Review): Pages 24-25, Lines 495-501**  TRDS 22 (**Completion date): NA**  TRDS 23 (**Summary Results): NA**  TRDS 24 **(IPD sharing statement): Page 25, Lines 516-519** | TRDS 3: NA; There are no secondary identifying numbers  TRDS 6: NA; There are no secondary sponsors  TRDS 22: NA; The study has not been completed  TRDS 23: NA; There are no results yet to summarize |
| Protocol version | [#3](https://www.goodreports.org/reporting-checklists/spirit/info/#3) | Date and version identifier | NA | This is the first published version, upon the date of publication. Version numbers for amendments and minor updates are tracked in the statistical analysis plan, which is available upon request, and updated in the clinical trials registry. |
| Funding | [#4](https://www.goodreports.org/reporting-checklists/spirit/info/#4) | Sources and types of financial, material, and other support | Page 25, Lines 503-506 |  |
| Roles and responsibilities: contributorship | [#5a](https://www.goodreports.org/reporting-checklists/spirit/info/#5a) | Names, affiliations, and roles of protocol contributors | Page 26, Lines 512-514 |  |
| Roles and responsibilities: sponsor contact information | [#5b](https://www.goodreports.org/reporting-checklists/spirit/info/#5b) | Name and contact information for the trial sponsor | Page 1, Lines 27-28; additional information at the link provided on Page 2, Line 54 |  |
| Roles and responsibilities: sponsor and funder | [#5c](https://www.goodreports.org/reporting-checklists/spirit/info/#5c) | Role of study sponsor and funders, if any, in study design; collection, management, analysis, and interpretation of data; writing of the report; and the decision to submit the report for publication, including whether they will have ultimate authority over any of these activities | Page 25, Lines 505-506 |  |
| Roles and responsibilities: committees | [#5d](https://www.goodreports.org/reporting-checklists/spirit/info/#5d) | Composition, roles, and responsibilities of the coordinating centre, steering committee, endpoint adjudication committee, data management team, and other individuals or groups overseeing the trial, if applicable (see Item 21a for data monitoring committee) | Page 24 Lines 482-487 |  |
| **Introduction** |  |  |  |  |
| Background and rationale | [#6a](https://www.goodreports.org/reporting-checklists/spirit/info/#6a) | Description of research question and justification for undertaking the trial, including summary of relevant studies (published and unpublished) examining benefits and harms for each intervention | Pages 3-5; Lines 60-119 |  |
| Background and rationale: choice of comparators | [#6b](https://www.goodreports.org/reporting-checklists/spirit/info/#6b) | Explanation for choice of comparators | Page 9, Lines 215-218 |  |
| Objectives | [#7](https://www.goodreports.org/reporting-checklists/spirit/info/#7) | Specific objectives or hypotheses | Page 5, Lines 128-132 |  |
| Trial design | [#8](https://www.goodreports.org/reporting-checklists/spirit/info/#8) | Description of trial design including type of trial (eg, parallel group, crossover, factorial, single group), allocation ratio, and framework (eg, superiority, equivalence, non-inferiority, exploratory) | Page 5, Line 124 |  |
| **Methods: Participants, interventions, and outcomes** | | | | |
| Study setting | [#9](https://www.goodreports.org/reporting-checklists/spirit/info/#9) | Description of study settings (eg, community clinic, academic hospital) and list of countries where data will be collected. Reference to where list of study sites can be obtained | Pages 6-7, Lines 147-155 |  |
| Eligibility criteria | [#10](https://www.goodreports.org/reporting-checklists/spirit/info/#10) | Inclusion and exclusion criteria for participants. If applicable, eligibility criteria for study centres and individuals who will perform the interventions (eg, surgeons, psychotherapists) | Pages 7-8, Lines 156-181 |  |
| Interventions: description | [#11a](https://www.goodreports.org/reporting-checklists/spirit/info/#11a) | Interventions for each group with sufficient detail to allow replication, including how and when they will be administered | Pages 9-10, Lines 210-235 |  |
| Interventions: modifications | [#11b](https://www.goodreports.org/reporting-checklists/spirit/info/#11b) | Criteria for discontinuing or modifying allocated interventions for a given trial participant (eg, drug dose change in response to harms, participant request, or improving / worsening disease) | Page 23, Lines 459-465; Page 10, Lines 233-234 |  |
| Interventions: adherance | [#11c](https://www.goodreports.org/reporting-checklists/spirit/info/#11c) | Strategies to improve adherence to intervention protocols, and any procedures for monitoring adherence (eg, drug tablet return; laboratory tests) | Page 15, Lines 260-264 |  |
| Interventions: concomitant care | [#11d](https://www.goodreports.org/reporting-checklists/spirit/info/#11d) | Relevant concomitant care and interventions that are permitted or prohibited during the trial | Page 10, Lines 234-235 |  |
| Outcomes | [#12](https://www.goodreports.org/reporting-checklists/spirit/info/#12) | Primary, secondary, and other outcomes, including the specific measurement variable (eg, systolic blood pressure), analysis metric (eg, change from baseline, final value, time to event), method of aggregation (eg, median, proportion), and time point for each outcome. Explanation of the clinical relevance of chosen efficacy and harm outcomes is strongly recommended | **Pages 11-14 (Table 1)** |  |
| Participant timeline | [#13](https://www.goodreports.org/reporting-checklists/spirit/info/#13) | Time schedule of enrolment, interventions (including any run-ins and washouts), assessments, and visits for participants. A schematic diagram is highly recommended (see Figure) | Page 16, Figure 4 |  |
| Sample size | [#14](https://www.goodreports.org/reporting-checklists/spirit/info/#14) | Estimated number of participants needed to achieve study objectives and how it was determined, including clinical and statistical assumptions supporting any sample size calculations | Pages 8-9, Lines 183-202 |  |
| Recruitment | [#15](https://www.goodreports.org/reporting-checklists/spirit/info/#15) | Strategies for achieving adequate participant enrolment to reach target sample size | Page 9, Line 208 |  |
| **Methods: Assignment of interventions (for controlled trials)** | | | | |
| Allocation: sequence generation | [#16a](https://www.goodreports.org/reporting-checklists/spirit/info/#16a) | Method of generating the allocation sequence (eg, computer-generated random numbers), and list of any factors for stratification. To reduce predictability of a random sequence, details of any planned restriction (eg, blocking) should be provided in a separate document that is unavailable to those who enrol participants or assign interventions | Pages 15-16, Lines 274-287 |  |
| Allocation concealment mechanism | [#16b](https://www.goodreports.org/reporting-checklists/spirit/info/#16b) | Mechanism of implementing the allocation sequence (eg, central telephone; sequentially numbered, opaque, sealed envelopes), describing any steps to conceal the sequence until interventions are assigned | NA | There is no concealment from individual participants. |
| Allocation: implementation | [#16c](https://www.goodreports.org/reporting-checklists/spirit/info/#16c) | Who will generate the allocation sequence, who will enrol participants, and who will assign participants to interventions | Page 15, Lines 280-282 |  |
| Blinding (masking) | [#17a](https://www.goodreports.org/reporting-checklists/spirit/info/#17a) | Who will be blinded after assignment to interventions (eg, trial participants, care providers, outcome assessors, data analysts), and how | Pages 21-22, Lines 415-420 |  |
| Blinding (masking): emergency unblinding | [#17b](https://www.goodreports.org/reporting-checklists/spirit/info/#17b) | If blinded, circumstances under which unblinding is permissible, and procedure for revealing a participant’s allocated intervention during the trial | NA | This is at the discretion of the DSMC. |
| **Methods: Data collection, management, and analysis** | | | | |
| Data collection plan | [#18a](https://www.goodreports.org/reporting-checklists/spirit/info/#18a) | Plans for assessment and collection of outcome, baseline, and other trial data, including any related processes to promote data quality (eg, duplicate measurements, training of assessors) and a description of study instruments (eg, questionnaires, laboratory tests) along with their reliability and validity, if known. Reference to where data collection forms can be found, if not in the protocol | Pages 16-21; Page 25, Lines 516-519 |  |
| Data collection plan: retention | [#18b](https://www.goodreports.org/reporting-checklists/spirit/info/#18b) | Plans to promote participant retention and complete follow-up, including list of any outcome data to be collected for participants who discontinue or deviate from intervention protocols | Pages 23-24, Lines 467-480 |  |
| Data management | [#19](https://www.goodreports.org/reporting-checklists/spirit/info/#19) | Plans for data entry, coding, security, and storage, including any related processes to promote data quality (eg, double data entry; range checks for data values). Reference to where details of data management procedures can be found, if not in the protocol | Page 22, Lines 422-431; Page 25, Line 518 |  |
| Statistics: outcomes | [#20a](https://www.goodreports.org/reporting-checklists/spirit/info/#20a) | Statistical methods for analysing primary and secondary outcomes. Reference to where other details of the statistical analysis plan can be found, if not in the protocol | Pages 22-23, Lines 437-457; Page 25, Lines 518 |  |
| Statistics: additional analyses | [#20b](https://www.goodreports.org/reporting-checklists/spirit/info/#20b) | Methods for any additional analyses (eg, subgroup and adjusted analyses) | Page 23, Lines 452-457 |  |
| Statistics: analysis population and missing data | [#20c](https://www.goodreports.org/reporting-checklists/spirit/info/#20c) | Definition of analysis population relating to protocol non-adherence (eg, as randomised analysis), and any statistical methods to handle missing data (eg, multiple imputation) | Pages 23-24, Lines 457-480 |  |
| **Methods: Monitoring** | | | | |
| Data monitoring: formal committee | [#21a](https://www.goodreports.org/reporting-checklists/spirit/info/#21a) | Composition of data monitoring committee (DMC); summary of its role and reporting structure; statement of whether it is independent from the sponsor and competing interests; and reference to where further details about its charter can be found, if not in the protocol. Alternatively, an explanation of why a DMC is not needed | Page 24, Lines 482-487 |  |
| Data monitoring: interim analysis | [#21b](https://www.goodreports.org/reporting-checklists/spirit/info/#21b) | Description of any interim analyses and stopping guidelines, including who will have access to these interim results and make the final decision to terminate the trial | Page 23, Lines 459-465 |  |
| Harms | [#22](https://www.goodreports.org/reporting-checklists/spirit/info/#22) | Plans for collecting, assessing, reporting, and managing solicited and spontaneously reported adverse events and other unintended effects of trial interventions or trial conduct | Page 22, Lines 425-426 |  |
| Auditing | [#23](https://www.goodreports.org/reporting-checklists/spirit/info/#23) | Frequency and procedures for auditing trial conduct, if any, and whether the process will be independent from investigators and the sponsor | Page 24, Lines 484-485 |  |
| **Ethics and dissemination** | | | | |
| Research ethics approval | [#24](https://www.goodreports.org/reporting-checklists/spirit/info/#24) | Plans for seeking research ethics committee / institutional review board (REC / IRB) approval | Pages 24-25, Lines 495-501 |  |
| Protocol amendments | [#25](https://www.goodreports.org/reporting-checklists/spirit/info/#25) | Plans for communicating important protocol modifications (eg, changes to eligibility criteria, outcomes, analyses) to relevant parties (eg, investigators, REC / IRBs, trial participants, trial registries, journals, regulators) | NA | Protocol amendments are approved by IRBs and updated in the clinical trials registry. |
| Consent or assent | [#26a](https://www.goodreports.org/reporting-checklists/spirit/info/#26a) | Who will obtain informed consent or assent from potential trial participants or authorised surrogates, and how (see Item 32) | Page 9, Line 205 |  |
| Consent or assent: ancillary studies | [#26b](https://www.goodreports.org/reporting-checklists/spirit/info/#26b) | Additional consent provisions for collection and use of participant data and biological specimens in ancillary studies, if applicable | NA | Ancillary studies are not planned. However, consent forms include use of deidentified data and specimens for future studies. |
| Confidentiality | [#27](https://www.goodreports.org/reporting-checklists/spirit/info/#27) | How personal information about potential and enrolled participants will be collected, shared, and maintained in order to protect confidentiality before, during, and after the trial | Page 22, Lines 433-435 |  |
| Declaration of interests | [#28](https://www.goodreports.org/reporting-checklists/spirit/info/#28) | Financial and other competing interests for principal investigators for the overall trial and each study site | Page 25, Lines 525-528 |  |
| Data access | [#29](https://www.goodreports.org/reporting-checklists/spirit/info/#29) | Statement of who will have access to the final trial dataset, and disclosure of contractual agreements that limit such access for investigators | Page 25, Lines 516-519 |  |
| Ancillary and post trial care | [#30](https://www.goodreports.org/reporting-checklists/spirit/info/#30) | Provisions, if any, for ancillary and post-trial care, and for compensation to those who suffer harm from trial participation | NA | No ancillary or post-trial care is anticipated. |
| Dissemination policy: trial results | [#31a](https://www.goodreports.org/reporting-checklists/spirit/info/#31a) | Plans for investigators and sponsor to communicate trial results to participants, healthcare professionals, the public, and other relevant groups (eg, via publication, reporting in results databases, or other data sharing arrangements), including any publication restrictions | Page 25, Lines 519-520 |  |
| Dissemination policy: authorship | [#31b](https://www.goodreports.org/reporting-checklists/spirit/info/#31b) | Authorship eligibility guidelines and any intended use of professional writers | NA | Authorship will follow ICMJE guidelines. |
| Dissemination policy: reproducible research | [#31c](https://www.goodreports.org/reporting-checklists/spirit/info/#31c) | Plans, if any, for granting public access to the full protocol, participant-level dataset, and statistical code | Page 25, Lines 516-520 |  |
| **Appendices** | | | | |
| Informed consent materials | [#32](https://www.goodreports.org/reporting-checklists/spirit/info/#32) | Model consent form and other related documentation given to participants and authorised surrogates | NA | Materials will be made available upon publication of results. |
| Biological specimens | [#33](https://www.goodreports.org/reporting-checklists/spirit/info/#33) | Plans for collection, laboratory evaluation, and storage of biological specimens for genetic or molecular analysis in the current trial and for future use in ancillary studies, if applicable | Pages 19-21, Lines 356-413 |  |

It is strongly recommended that this checklist be read in conjunction with the SPIRIT 2013 Explanation & Elaboration for important clarification on the items. Amendments to the protocol should be tracked and dated. The SPIRIT checklist is copyrighted by the SPIRIT Group under the Creative Commons “[Attribution-NonCommercial-NoDerivs 3.0 Unported](http://www.creativecommons.org/licenses/by-nc-nd/3.0/)” license. This checklist can be completed online using https://www.goodreports.org/, a tool made by the EQUATOR Network in collaboration with Penelope.ai
